# Electroencephalographic (EEG) Changes Accompanying Normal Breathing of Concentrated Oxygen (Hyperoxic Ventilation) by Healthy Adults: A Systematic Review

**DOI:** 10.1101/2024.09.16.24313766

**Authors:** Lachlan D. Barnes, Luke E. Hallum, Xavier C.E. Vrijdag

**Affiliations:** Department of Anaesthesiology, University of Auckland; Department of Mechanical Engineering, University of Auckland

## Abstract

**Introduction:** Divers often increase their fraction of inspired oxygen (FiO_2_) to decrease their risk of decompression sickness. However, breathing concentrated oxygen can cause hyperoxia, and central nervous system oxygen toxicity (CNS-OT). This study aims to review the literature describing hyperoxic ventilation’s effect on the electroencephalogram (EEG), thus exploring the potential for real-time detection of impending CNS-OT seizure.

**Methods:** We searched Medline, Embase, Scopus, and Web of Science for articles that reported EEG measures accompanying hyperoxic ventilation (FiO_2_ = 1.0) in healthy participants. We included peer-reviewed journal articles, books, and government reports with no language or date restrictions. Randomised controlled trials and cross-over studies were included; case reports were excluded. We used the Newcastle-Ottawa scale to evaluate evidence quality.

**Results:** Our search strategy returned 1025 unique abstracts; we analysed the full text of 40 articles; 22 articles (16 studies) were included for review. Study cohorts were typically small, and comprised of male non-divers. We discovered a variety of EEG analysis methods: studies performed spectral analysis (*n* = 12), the analysis of sensory-evoked potentials (*n* = 4), connectivity/complexity analysis (*n* = 3), source localization (*n* = 1), and expert qualitative analyses (*n* = 4). Studies of severe exposures (long duration at hyperbaric pressure) typically reported qualitative measures, and studies of mild exposures typically reported quantitative measures.

**Conclusions:** There is a need for a large randomised controlled trial (RCT) reporting quantitative measures to better understand hyperoxic ventilation’s effect on EEG, thus enabling the development of real-time monitoring of CNS-OT risk.

## Introduction

Diver performance and safety is affected by the physical and chemical properties of the gas mixtures breathed.^1^ For example, technical divers working in shallow water (< 30 meters) will often breathe enriched (nitrox) gas mixtures;^2^ this, as compared to breathing normal air, reduces the severity of diving-related medical conditions such as decompression sickness.^3^ However, breathing high concentrations of oxygen (“hyperoxic ventilation”) and increased pressure promotes hyperoxia, the physiological state wherein oxygen levels in blood and tissue are abnormally high. This, in turn, increases the risk of central nervous system oxygen toxicity (CNS-OT), that is, poisoning that occurs due to exposure to elevated partial pressures of inspired oxygen.^4^ Several factors can promote hyperoxia, including the oxygen concentration of the breathed gas, the duration of exposure to this gas, barometric pressure, hypercapnia, and physical exertion.^5–7^ At present, the only way for divers to prevent CNS-OT is to choose conservative oxygen exposures.

CNS-OT can be mild to severe.^8^ Signs and symptoms of mild and moderate disease include tunnel vision, tinnitus, nausea, lip twitching, irritability, and dizziness.^9^ Severe symptoms involves seizures, convulsions, and unconsciousness;^10^ an unconscious diver is at high risk of losing their mouthpiece, and subsequent fatality. CNS-OT symptomatology is highly variable both between divers, and within a diver between dives.^7^ Often, a diver has no warning of impending seizure.^5^

CNS-OT seizures have been shown to alter EEG recordings.^11–13^ However, the precise nature of these EEG alterations is unclear. It is also unclear whether, in CNS-OT, EEG alterations occur abruptly or, rather, emerge gradually. The gradual emergence of EEG alterations could be used to predict an impending seizure, and therefore be useful in real-time monitoring of seizure risk during dives, or in clinical decision support. The aim of this systematic review is to collect EEG alterations known to be associated with hyperoxic ventilation and, therefore, identify those alterations potentially useful in predicting CNS-OT onset.

## Methods

### SEARCH STRATEGY

We conducted a systematic search using Medline, Embase, Scopus, and Web of Science (date of last search: 16 February 2024). We searched for entries labelled with the MeSH Headings “Electroencephalography” or “Electroencephalography phase synchonization”, or entries containing “electroencephalogram”, or synonyms thereof, anywhere in the title, abstract, or keywords. We limited these results to those with the MeSH Headings “Oxygen”, “Hyperbaric Oxygenation”, or “Hyperoxia”, or those containing the terms “hyperoxia”, “hyperoxemia”, or those with “oxygen” within two words of “hyperbaric” or “pressure”. Next, these results were limited to those labelled with the MeSH Heading “Diving”, or containing the terms “diving”, “diver”, “divers”, “hyperbaric”, or “normobaric”. Finally, we restricted these results to human studies. We imposed no restrictions on language or publication date; we translated non-English articles for screening and review. Our search strategy is fully specified in Appendix A. We drafted the search strategy with assistance from the University of Auckland librarian, and validated the strategy against five articles identified as matching the review protocol.^12, 14–17^ After developing the Medline search strategy, we translated it into formats compatible with the other three databases (Appendix A).

### SELECTION PROCESS

We imported search results from Medline, Embase, Scopus, and Web of Science into Covidence reference management software (Veritas Health Innovation Ltd., Melbourne, Australia; available at https://www.covidence.org) for deduplication, review, and data extraction. To guide our methods, we used the PRISMA statement.^18^ The authors (LB, LH, XV) screened titles and abstracts of articles discovered by the search; every title and abstract was screened independently by at least two authors. We excluded articles involving paediatric subjects, animals, chronic exposure scenarios, or patient cohorts (i.e., studies of participants with pre-existing medical conditions). We resolved any disputes regarding inclusion or exclusion through discussion.

We obtained full-text versions of all articles deemed relevant. After reviewing the full text, we excluded articles that either lacked primary data, used exposures other than pure oxygen (i.e., FiO_2_ < 1.0), or failed to report EEG outcomes. We then screened the citations within the included articles, adding relevant references to the full-text review. Next, we conducted citation searches on these articles using two tools: ResearchRabbit (ResearchRabbit, United States of America; available at https://www.researchrabbit.ai), and PaperFetcher.^19^ Both tools used the list of included references to identify additional relevant papers. ResearchRabbit generates a list of articles related to the supplied articles (they do not specify their methodology). PaperFetcher employs both forward and backward citation searches to compile a list of relevant articles. Forward citation searches find all articles that have cited the references, while backward citation searches find all articles cited by any of the articles in the reference list. All discoveries from this citation search underwent title and abstract screening before any full-text review.

### DATA EXTRACTION

Authors LB and LH extracted data using a custom data extraction form (Appendix B). The data extracted included study design, hyperoxic ventilation exposure, participant demographics, and quantitative and qualitative EEG results. Given the diversity in quantitative data and experimental designs, a formal meta-analysis was not feasible.

Reviewers LB and LH employed the Newcastle-Ottawa scale (NOS) for assessing the quality of studies^20^ (Appendix B). This scale ranges from zero to nine, where zero (nine) indicates the worst (best) possible quality. The NOS evaluates three aspects of studies: (1) cohort selection, scoring up to 4 points for representativeness; (2) comparability between study groups, scoring up to 2 points for effective control of confounding variables (e.g., age and gender); and, (3) integrity of outcome assessments, scoring up to 3 points based on blind evaluation, sufficient outcome manifestation time, and thorough follow-up. The overall score is converted to a measure of quality using the Agency for Healthcare Research Quality guidelines^21^ as follows:

- **Good quality:** Requires 3 to 4 points in selection, and 1 to 2 points in comparability, and 2 to 3 points in outcome assessment.
- **Fair quality:** Requires 2 points in selection, and 1 to 2 points in comparability, and 2 to 3 points in outcome assessment.
- **Poor quality:** 0 or 1 points in selection, 0 in comparability, 0 or 1 in outcome assessment.

## Results

### INCLUDED STUDIES

Our search across the four databases yielded 1,115 articles. Additionally, citation searching contributed 403 articles, and manual citation searching added seven more. After deduplication, we screened the titles and abstracts of 1,025 articles. We read the full text of 40 articles; we included 22 of these articles in this review. These 22 articles reported 16 studies (i.e., several articles reported the same primary data). We illustrate the article selection process in Figure 1.

**FIGURE 1:**
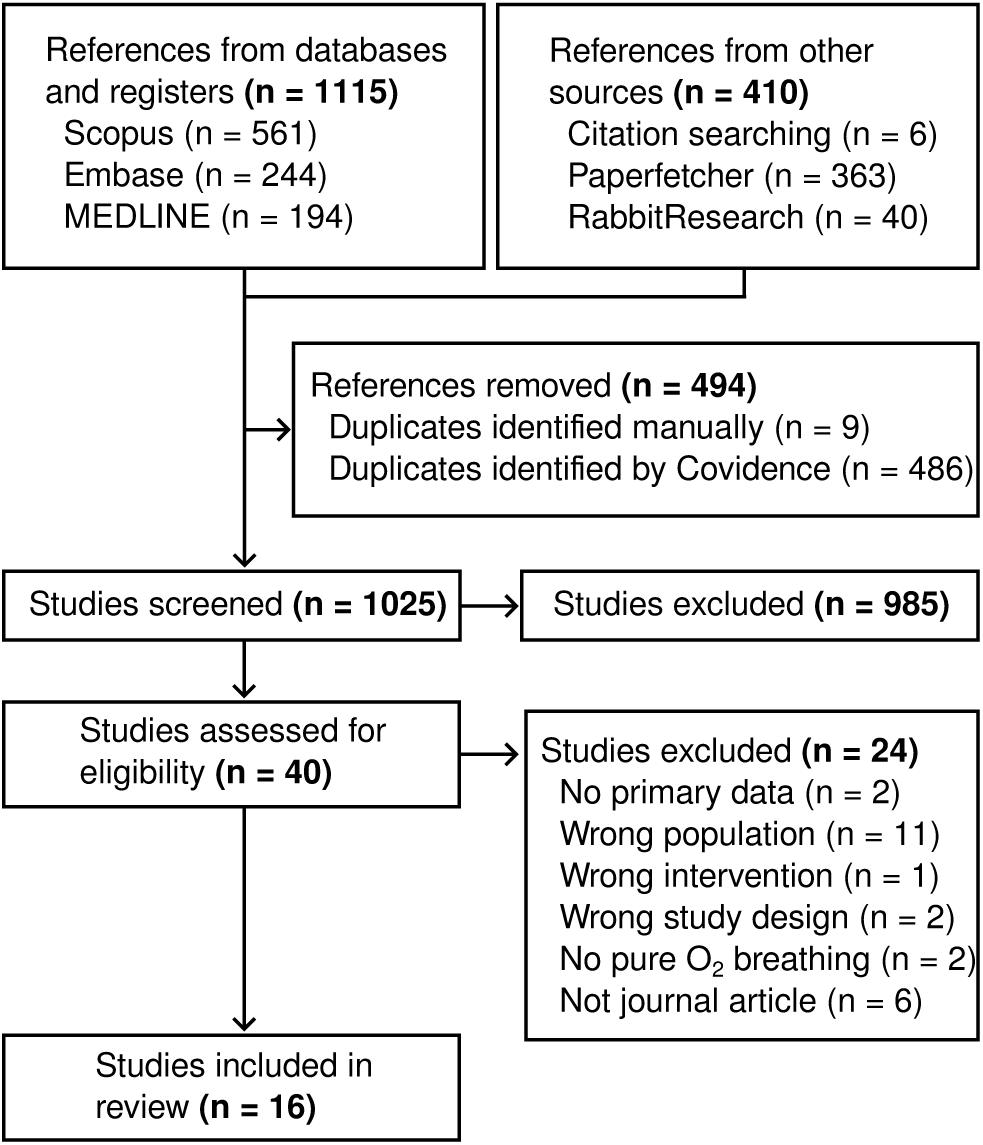
Flow diagram of article selection.

Our process selected one randomized control trial;^22^ the remaining studies were non-randomized and/or non-controlled. Most studies used a cross-over design. Of the 16 studies included in this review, we graded seven as “good” quality, none as “fair”, and nine as “poor”. All “poor” studies failed to control for age, sex, marital status, or other factors, and therefore failed to score “comparability” points on the NOS (Table 1).

**TABLE 1:** We used the Newcastle-Ottawa scale to assess the quality of the 16 studies included in our review. The table shows the count of studies that received each score for each aspect of the Newcastle-Ottawa scale.

| Score | Selection (max. 4 points) | Comparability (max. 2 points) | Assessed outcome (max. 3 points) |
| --- | --- | --- | --- |
| ≥ 3 points | 15 |  | 16 |
| 2 points | 1 | 0 | 0 |
| 1 point | 0 | 7 | 0 |
| 0 points | 0 | 9 | 0 |

### STUDY PARTICIPANTS

In most of the studies included in our review, the cohort size was less than 15. The study with the largest cohort included 39 participants, however, this study used normobaric, not hyperbaric, exposure, meaning that the risk of CNS-OT was much reduced.^15^ The largest hyperbaric study involved 34 participants across five separate exposures.^13^ Typically, experimental participants were male; overall, 80% of participants were male; only two studies used majority-female cohorts.^23, 24^ Most participants were between 30 and 39 years old; the youngest participant was 18 years,^23^ while the oldest, was 81 years.^25^ Cohort demographics are tabulated in Table 2.

**TABLE 2:** Study participant characteristics.

| Parameter | Number of studies<br>(n=16) |
| --- | --- |
| **Number of participants** |  |
| < 10 | 2 |
| 10-15 | 7 |
| 16-20 | 1 |
| > 20 | 6 |
| Not specified | 2 |
| **Male percentage** |  |
| 100% | 7 |
| 50-99% | 3 |
| < 50% | 2 |
| Not specified | 4 |
| **Average age (years)** |  |
| 18-19 | 0 |
| 20-29 | 3 |
| 30-39 | 5 |
| 40-49 | 1 |
| 50-59 | 0 |
| Not specified | 7 |
| **Characteristic** |  |
| Healthy adult volunteers | 10 |
| Healthy adult volunteers with some diving experience (< 1 year) | 4 |
| Healthy adult volunteers with moderate diving experience (< 3 years) | 0 |
| Healthy adult volunteers with significant diving experience (3+ years) | 2 |
| **Participants who had conducted an oxygen tolerance test** |  |
| 100% | 1 |
| 50-99% | 0 |

Participants were healthy adult volunteers, and most had no reported diving experience (10 out of 16 studies). Four studies involved individuals with some diving experience (either unspecified or less than one year),^11, 12, 16, 26^ while two studies recruited participants with significant diving experience (three or more years).^14, 27^ Most studies failed to report if participants had prior hyperoxic ventilation exposures. In two studies, some participants had undergone oxygen tolerance tests,^12, 14^ and in one study, four individuals had prior hyperoxic episodes.^14^

### INTERVENTION

Among the 16 studies included in our review, four included multiple oxygen exposures (all were FiO_2_ = 1.0) at different durations and/or hyperbaric pressures.^13, 16, 27, 28^ One study featured five exposures,^13^ two studies each included three exposures,^16, 27^ and one study included two exposures.^28^ Of these 25 exposures, we classified seven as mild, four as moderate, and 14 as severe. We defined mild exposures as those that occurred at normobaric pressure. Moderate exposures were hyperbaric, but did not exceed the safety guidelines set by the US Navy Dive Manual Rev. 7A for single-depth oxygen dives.^29^ Severe exposures were hyperbaric exposures that surpassed these guidelines.

There were three “outlier” exposures which far exceeded the US Navy Diving Manual Guidelines.^11, 13, 28^ Of these three studies, the data collection for two occurred during wartime (World War II).^11, 13^ The greatest hyperbaric exposure was at 472.2 kPa (duration not specified),^11^ and the greatest exposure duration was 24 hours (at 101.3 kPa).^13^ Figure 2 illustrates the range of interventions described by the studies included in our review.

**FIGURE 2:**
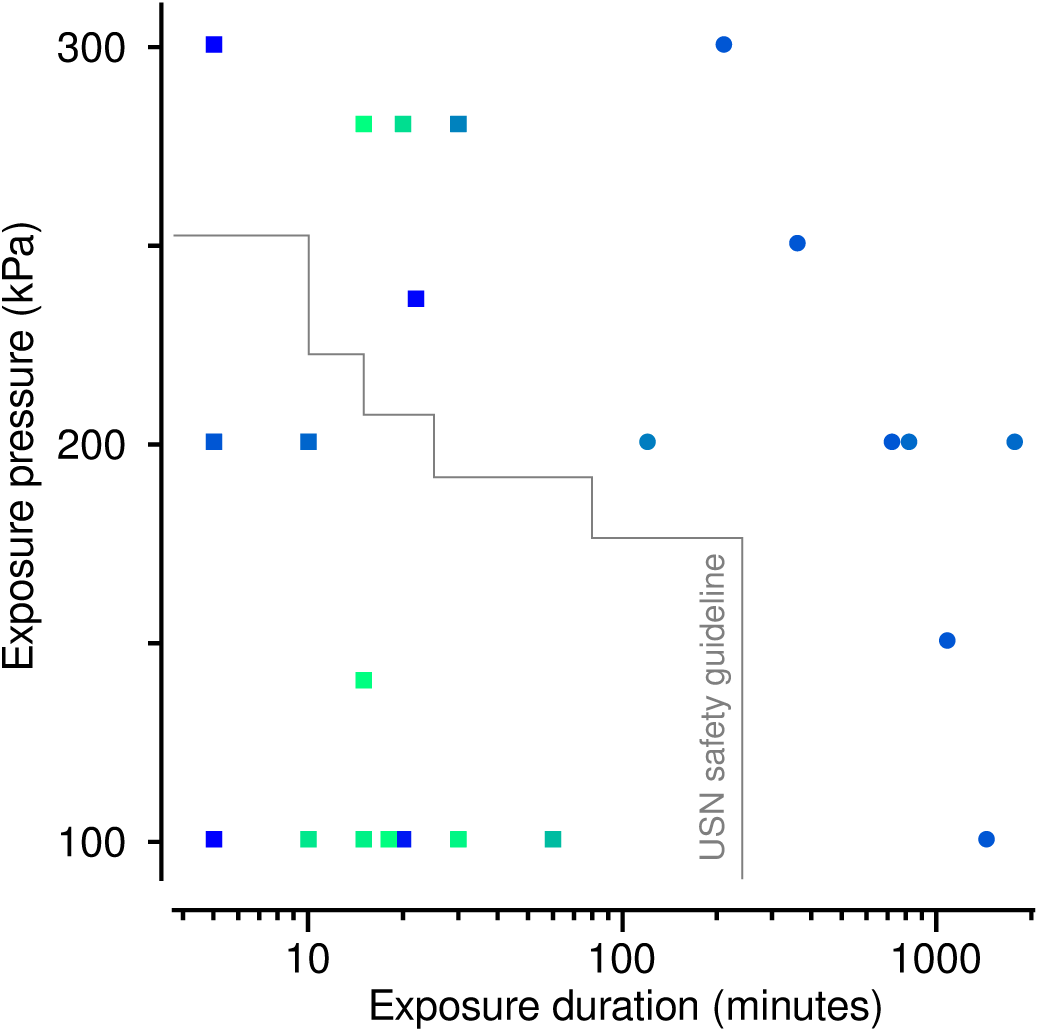
Hyperoxic ventilation exposures described by studies included in our review. The grey line shows the US Navy (USN) Diving Manual Guidelines.^27^ Each symbol represents an exposure. Squares represent studies reporting quantitative measures, whereas circles represent studies reporting only qualitative measures. Colour represents the year of study publication: green (blue) symbols show recent (earlier) studies.

### INDUCTION OF HYPEROXIA

In the 16 studies reviewed, all exposures used either normobaric conditions or used a hyperbaric chamber to create hyperbaric conditions. Most of these studies delivered oxygen via a face mask.^12–14, 22–24, 27, 28, 30–33^ However, there were exceptions: a Siebe Gorman “salvus “ apparatus;^11^ a clear plastic tent;^25^ a mouthpiece with a two-way valve,^15^ and a mouthpiece with an on-demand valve.^16^

### EEG FREQUENCY ANALYSIS

Of the 16 studies reviewed, 12 reported using quantitative spectral analysis of EEG. These results are summarized in Table 3.

**Table 3:** Overview of EEG frequency-band power alterations with hyperoxic ventilation. Here, we have excluded studies that reported no relationship between power alterations and exposures. Increases in power are indicated by ↑, decreased by ↓, and no change by ↔. Statistically significant results are indicated by *, while results without any reported statistical testing are underlined.

| Study | Exposure | Duration<br>(minutes) | Exposure<br>(kPa) | Alpha | Beta | Delta | Theta |
| --- | --- | --- | --- | --- | --- | --- | --- |
| Pastena<sup>14</sup> | Severe | 20 | 283.7 | ↑* | ↑* | ↓* | ↓* |
| Visser<sup>12</sup> | Severe | 30 | 283.7 | ↑ (upper<br>alpha)<br>↓ (lower<br>alpha) | ↔ | ↔ | ↔ |
| Donald<sup>11</sup> | Severe | Not<br>specified | 472.2 | ↓ | ↑ | ↔ | ↑ |
| Litscher<sup>33</sup> | Moderate | 10 | 202.7 | ↑ | ↔ | ↓ | ↔ |
| Kimmo<sup>30</sup> | Mild | 60 | 101.3 | ↓ | ↔ | ↑ | ↔ |
| Damato<sup>22</sup> | Mild | 30 | 101.3 | ↑ | ↔ | ↔ | ↔ |
| Kizuk<sup>23</sup> | Mild | 15 | 101.3 | ↓* | ↓* | ↑* | ↑* |
| Sheng<sup>15</sup> | Mild | 10 | 101.3 | ↓* | ↓* | ↔ | ↔ |

#### Alpha waves

Studies relating alpha-band power to hyperoxic ventilation produced mixed results. At normobaric pressure, four studies reported change in alpha-band power.^15, 22, 23, 30^ Sheng and colleagues reported a 15.6% reduction in alpha-band power during rest (hyperoxic ventilation versus normoxic ventilation) and a 7.7% reduction in alpha-band power while measuring visual evoked responses.^15^ Similarly, Kizuk and colleagues^23^ observed a reliable decrease in alpha-band power, but only when participants’ eyes were open. Kimmo et al. also reported a decrease in alpha-band power, but this finding was a trend only.^30^ In contrast, Damato et al. reported increased alpha-band power at normobaric pressure.^22^

At hyperbaric pressure, several studies reported change in alpha-band power, likewise with mixed results.^11, 12, 14, 33^ Litscher and colleagues, reported an increase in alpha-band power (hyperoxic ventilation versus normoxic ventilation), however, this change was not statistically significant.^33^ Similarly, Pastena et al. reported an increase in alpha-band power at posterior recording sites.^14^ In contrast, some studies have reported decreased alpha-band power.^11^ However, this conclusion was made using visual analysis (i.e., qualitative analysis) of the recorded EEG, and no statistical analysis was provided. Visser et al., observed an increase in upper-frequency alpha-band power and a decrease in lower-frequency alpha-band power during hyperoxic ventilation (compared to normoxic).^12^

#### Beta waves

Studies relating beta-band power to hyperoxic ventilation also produced mixed results. For normobaric exposure, four studies concluded an association between hyperoxia-ventilation and beta-wave activity.^11, 14, 15, 23^ Two of these studies reported decreased beta-band power (hyperoxic ventilation versus normoxic ventilation).^15, 23^ In one case, this decrease was only observed when participants’ eyes were open.^23^ Sheng and colleagues observed this change only during the measurement of visual evoked responses.^15^ For hyperbaric exposure, two studies reported an increase in beta-band power linked to hyperoxia.^11, 34^ Pastena and colleagues found that this decrease was limited to the upper beta band (13 to 30 Hz) and primarily observed over temporal cortext.^14^ A similar result was reported by Donald et al., but this change in the beta-band power was observed only in the 25 to 32 Hz range.^11^

#### Theta waves

Studies relating theta-band power to hyperoxic ventilation, again, produced mixed results. For normobaric exposure, Kizuk et al. reported an increase in theta-band power (hyperoxic ventilation versus normoxic ventilation) during their eyes-closed condition – a change which was focused around the right-frontal region.^23^ For hyperbaric exposure, Pastena et al., observed a decrease in theta-band power; this change persisted throughout hyperoxic ventilation, and was focused over the parental region.^14^ In contrast, Donald et al. reported an increase in theta-band power.^11^

#### Delta Waves

Results relating delta-band power to hyperoxic ventilation were, too, mixed. For normobaric exposure, two studies identified effects of hyperoxic ventilation on delta waves.^23, 30^ Kizuk and colleagues reported an increase at the right-posterior electrode sites while participants’ eyes were closed.^23^ Kimmo and colleagues reported an increase over frontal and temporal cortical regions.^29^ For hyperbaric exposure, two studies reported a decrease in delta-band power.^14, 33^ Pastena et al. reported statistically significant decreases at posterior electrodes;^14^ this decrease in delta-band power was accompanied by a simultaneous increase in alpha-band power at the same site. Litscher et al. described a decrease in delta-band power activity, however, this change was not statistically significant.^14, 33^

### OTHER EEG ANALYSIS TECHNIQUES

Among the 16 studies included in our review, spectral analysis was not the only technique used. Additional techniques fell into three broad categories: evoked potentials (reported by four studies),^15, 27, 31, 33^ connectivity/complexity analysis (reported by three),^16, 34, 35^ source localization (reported by one),^34, 35^ and qualitative analysis (reported by four).^11, 13, 28, 32^

#### Evoked potentials

Four studies measured evoked potentials during hyperoxic ventilation: one employed visual stimuli,^15^ another used auditory and somatosensory stimuli,^33^ a third applied auditory and visual stimuli,^26^ and the fourth used auditory stimuli.^27^ Two of these studies reported significant changes in evoked potential responses (hyperoxic versus normoxic ventilation).^15, 27^ Sheng and colleagues reported a delay in N1 and P2 components of the visual evoked potential (VEP), but found no change in VEP amplitude.^15^ In contrast, Bennett and colleagues reported a reduction in the amplitude of auditory evoked potentials (AEPs) (hyperoxic versus normoxic ventilation); the magnitude of this reduction increased with hyperbaric pressure.^27^ Litscher et al., noted a small change in brainstem evoked potentials, but this change was not statistically significant.^33^

#### Connectivity/complexity analysis

Two studies employed connectivity/complexity analysis techniques to understand the impact of hyperoxic ventilation on the brain.^16, 34^ Vrijdag and colleagues, found no significant change in connectivity (hyperoxic versus normoxic ventilation).^16^ However, a significant reduction in temporal complexity was reported. These researchers quantified temporal complexity by the entropy of the diagonal line-length probability distribution of the binarized cross-correlation matrices of consecutive time samples, indicating how variable the signal was over the medium time range (2-10 seconds).^16^ Storti et al. used multivariate autoregression to estimate the direction of information flow between cortical sites.^34^ They found an increase in connectivity from frontal to posterior cortical regions (hyperoxic versus normoxic ventilation), particularly within the alpha- and beta-band frequencies.

#### Source localisation

Pastena and colleagues used the source localisation technique to estimate the origin of EEG alterations associated with hyperoxic ventilation;^34, 35^ to do so they used sLORETA (standardized low-resolution brain electromagnetic tomography).^36^ They found when participants breathed pure oxygen at 283.7 kPa, there was a rapid and statistically significant reduction in delta- and theta-band sources in the posterior region of the brain. Simultaneously, there was an apparent increase in power in the alpha and lower-beta (12 to 18 Hz) bands which was localized to the posterior region.

#### Qualitative analysis

Four studies conducted qualitative analysis of EEG recordings.^11, 13, 28, 32^ In all cases, experts reviewed the electroencephalogram, but found no EEG patterns consistently associated with hyperoxic ventilation, nor any preceding CNS-OT seizure. In work by Donald and colleagues,^11^ fifteen participants from the Admiralty Experimental Diving Unit were ranked on oxygen tolerance. This ranking was based on multiple dives at 286.6, 379.3, and 471.9 kPa in dry conditions, and 255.7 kPa in wet conditions. Dry EEG recordings were then classified as normal, abnormal or doubtful based on two independent opinions (the criteria used for these classifications was not reported). Of these, five participants had normal EEGs, seven had doubtful EEGs, and three had abnormal recordings. Notably, the three participants with the highest oxygen tolerance exhibited normal EEGs; however, all participants who experienced convulsions during, or after, the intervention had either abnormal or doubtful EEGs. However, the third most oxygen tolerant participant, who initially had a normal EEG, convulsed after intervention.

#### EEG Changes Accompanying CNS-OT Seizures

Among the 16 studies included in our review, three documented EEG changes during CNS-OT seizures.^11–13^ Donald and colleagues^11^ observed that, in some cases, there were bursts of theta-band activity, with increasing voltage just before seizure. However, in other cases, they reported no observable change in cortical activity preceding seizure. In a study by Visser et al.,^12^ an experienced diver, who had passed an oxygen-tolerance test three years earlier, experience seizure at the end of his 30-minute exposure to hyperbaric hyperoxic ventilation (283.7 kPa). This diver’s breathing became irregular 135 seconds before seizure onset due to abdominal myoclonic jerks. Throughout the seizure, no lateralizing signs or epileptiform activities were visually detected. However, there was a noticeable increase in theta-wave activity, both isolated and in short bursts, alongside a slowing of the alpha rhythm. The power spectrum showed a mild increase, particularly in the theta and delta bands, about 3 to 4 minutes before the seizure onset. This change coincided with the initial clinical respiratory signs. The EEG patterns during the seizure were consistent with those typical of a tonic-clonic seizure.

In work by Lambertsen and colleagues, two participants were exposed to hyperoxic ventilation at 304.0 kPa.^13^ The first participant experienced seizure after three hours of exposure, showing typical tonic-clonic EEG patterns; this study reported no change in EEG prior to seizure onset.^13^ The second participant, after 2.5 hours, exhibited a 10-second flat (i.e., isoelectric) EEG period, accompanied by 20 seconds of hypotensive unconsciousness. Recovery was marked by a mild tonic-clonic seizure and 30 seconds of disorganized EEG activity, after which normal EEG activity resumed.

## Discussion

Our systematic search of the literature discovered 16 studies (22 articles) reporting electroencephalogram alterations (or a lack thereof) that accompany hyperoxic ventilation (FiO_2_ = 1.0) in healthy adults. We were surprised by the paucity of data on this topic; most studies were observational, designs were heterogeneous, and results were inconsistent. There appears to be a need for a large randomized, controlled trial on the cortical effects of hyperoxic ventilation on normal participants. Across studies, we found no consistent association between hyperoxic ventilation and alterations in the power spectrum of recorded EEG. Quantitative analyses of EEG recordings other than the spectral analysis show promise, but are limited: connectivity/complexity analysis may signal hyperoxia, but these results need independent replication; visual- and auditory-evoked potentials may also signal hyperoxia, but the analysis of evoked potentials in not applicable to real-time monitoring, which is our primary motivation for conducting this review.

The studies discovered by our search all involved small cohorts, typically comprised of male non-divers. Using the Newcastle-Ottawa scale, we judged most studies to be of poor quality, usually because they lacked comparability, that is, they failed to use experimental design or statistical analysis to control for confounding factors such as participant age and sex. Most studies did not produce results that achieved statistical significance; some studies reported statistical significance, but only for a subset of experimental conditions (e.g., only when analysing EEG recordings using a subset of EEG electrodes). Studies typically involved either quantitative analysis of EEG recordings during mild/moderate hyperoxic ventilation exposures (i.e., short-duration exposure at normo- or hyperbaric pressure), or qualitative analysis of EEG recordings during more severe exposures (i.e., long durations at hyperbaric pressure).

Our systematic searches discovered only studies conducted in dry experimental conditions during which participants were not exercising. No studies in wet experimental conditions is unsurprising due to the difficulties of measuring EEG underwater. Therefore, the results of these studies may have only limited applicability to diving. Donald and colleagues have previously reported that oxygen tolerance is diminished when participants are in wet, as opposed to dry, conditions.^37^ The current understanding is that immersion in water redistributes the body’s circulation, leading to increased regional cerebral blood flow (rCBF), which in turn reduces seizure onset latency.^38, 39^ Donald and colleagues confirmed earlier animal studies,^40^ finding that exercising participants show markedly diminished resistance to CNS-OT.^7, 37, 41^ Exercise appears to cause a build-up in carbon dioxide which interacts with nitric oxide production, thus resulting in increased CNS-OT susceptibility.^42^

Our systematic review discovered three studies that reported EEG alterations specifically associated with CNS-OT seizure.^11–13^ Results across these three studies were consistent in that EEG changes during seizure appear indistinguishable from tonic-clonic seizure. Two of these studies reported that seizure onset was preceded by a broadband power increase in general, and a theta-band power increase in particular.^11–13^ However, by contrast, Visser et al. reported no EEG alterations preceding seizure onset.^12^ Indeed, reports of CNS-OT seizure in the literature are few, and none of the studies reporting seizure is recent. It remains an open question whether there exist reliable EEG signs of impending seizure.^43^ We note that some animal studies indicate a relationship between rCBF, nitric oxide production and CNS oxygen toxicity. Demchenko and colleagues^44^ found hyperoxia leads to an increase in nitric oxide production, increased rCBF, causing surplus oxygen to be delivered to neuropil. This rise in rCBF preceded an increase in bursts of EEG activity, followed by seizure.^44^

## Conclusions

Can EEG be used to detect, in real time, an impending CNS-OT seizure? Our review revealed several shortcomings of the literature which, taken together, obviate a straightforward answer to that question. First, the 16 studies included in our review were small-cohort studies; cohort size likely contributed to the heterogeneous results we discovered. Second, none of these studies used an experimental set-up representative of diving. Water immersion and exercise necessarily accompany diving, and both are known to affect susceptibility to CNS-OT.^7^ However, none of the reviewed studies incorporated these factors into experimental design. Finally, most reviewed studies used either mild hyperoxic-ventilation exposures with quantitative EEG analysis, or moderate/severe exposures with qualitative analysis. Thus, these EEG findings have limited translational potential for real-time monitoring; mild exposure is unlikely to cause CNS-OT seizure, and qualitative expert analysis is difficult to implement by way of real-time software. We conclude that there is a need for further research into hyperoxic ventilation’s effect on EEG to help answer this open question.

## Data Availability

All data produced in the present work are contained in the manuscript.

No financial interests. No conflicts of interest.

## Acknowledgements

We thank Rayna Dewar, a librarian at the University of Auckland, for her expert guidance and support in creating the Medline search strategy. We also thank The National Archives for their assistance in conducting this research.

## Appendix A

### MEDLINE SEARCH STRATEGY

1. Electroencephalography/ or Electroencephalography phase synchronization/
2. (eeg or eegs or electroencephalogra*).mp.
3. 1 or 2
4. Oxygen/ or Hyperbaric Oxygenation/ or Hyperoxia/
5. (hyperoxi* or hyperox?em*).mp.
6. (oxygen adj2 hyperbar*).mp.
7. (oxygen adj2 pressu*).mp.
8. 4 or 5 or 6 or 7
9. Diving/
10. (Diving or diver or divers or hyperbaric or normobaric).mp.
11. 9 or 10
12. 3 and 8 and 11
13. limit 12 to humans

### EMBASE SEARCH STRATEGY

1. Electroencephalography/ Electroencephalography phase synchronization/ or Electroencephalogram/
2. (eeg or eegs or electroencephalogra*).mp.
3. 1 or 2
4. Oxygen/ or Hyperbaric Oxygenation/ or Hyperoxia/
5. (hyperoxi* or hyperox?em*).mp.
6. (oxygen adj2 hyperbar*).mp.
7. (oxygen adj2 pressu*).mp.
8. 4 or 5 or 6 or 7
9. Diving/
10. (Diving or diver or divers or hyperbaric or normobaric).mp.
11. 9 or 10
12. 3 and 8 and 11
13. limit 12 to humans

### SCOPUS SEARCH STRATEGY

1. INDEXTERMS(Electroencephalography)
2. TITLE-ABS-KEY(eeg) OR TITLE-ABS-KEY(eegs) OR TITLE-ABS-KEY(electroencephalogra*)
3. #1 OR #2
4. INDEXTERMS(Oxygen)
5. INDEXTERMS(Hyperbaric Oxygenation)
6. INDEXTERMS(Hyperoxia)
7. TITLE-ABS-KEY(hyperoxi*) OR TITLE-ABS-KEY(hyperox?em*)
8. TITLE-ABS-KEY(oxygen) W/2 TITLE-ABS-KEY(hyperbar*)
9. TITLE-ABS-KEY(oxygen) W/2 TITLE-ABS-KEY(pressu*)
10. #4 OR #5 OR #6 OR #7 OR #8 OR #9
11. INDEXTERMS(Diving)
12. TITLE-ABS-KEY(Diving) OR TITLE-ABS-KEY(diver) OR TITLE-ABS-KEY(divers) OR TITLE-ABS-KEY(hyperbaric) OR TITLE-ABS-KEY(normobaric)
13. #11 OR #12
14. #3 AND #10 AND #13
15. INDEXTERMS (animals) NOT INDEXTERMS (humans)
16. #14 AND NOT #15

### WEB OF SCIENCE SEARCH STRATEGY

1. KP=(electroencephalography)
2. TS =(eeg OR eegs OR electroencephalogra*)
3. #1 OR #2
4. KP=(Oxygen)
5. KP=(Hyperbaric Oxygenation)
6. KP=(Hyperoxia)
7. TS=(hyperoxi* OR hyperox*em*)
8. TS=(oxygen NEAR/2 hyperbar*)
9. TS=(oxygen NEAR/2 pressu*)
10. #4 OR #5 OR #6 OR #7 OR #8 OR #9
11. KP=(Diving)
12. TS=(Diving or diver or divers or hyperbaric or normobaric)
13. #11 OR #12
14. #3 AND #10 AND #13
15. #14 NOT TS=(animals NOT humans)

## Appendix B

### DATA EXTRACTION TEMPLATE

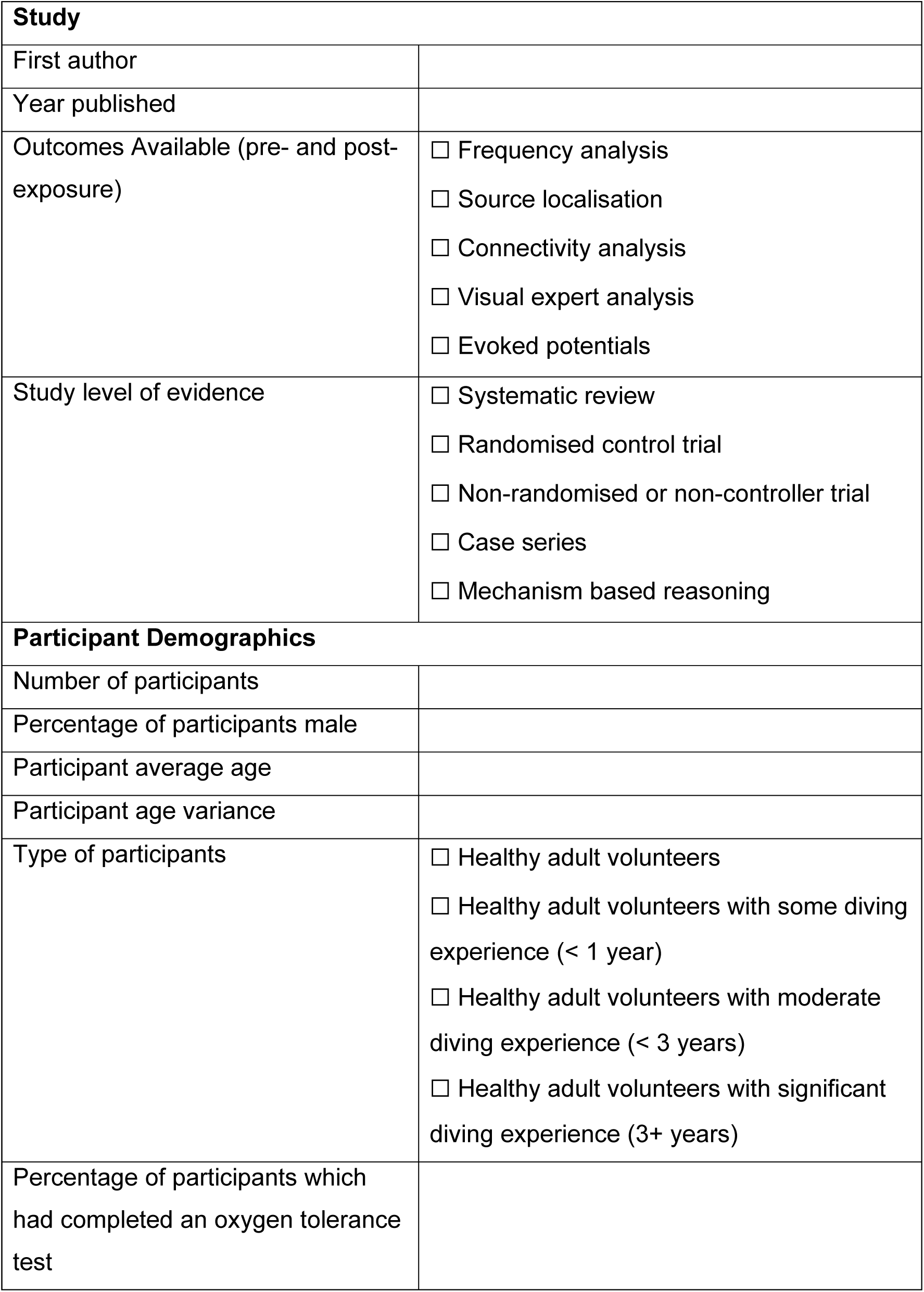
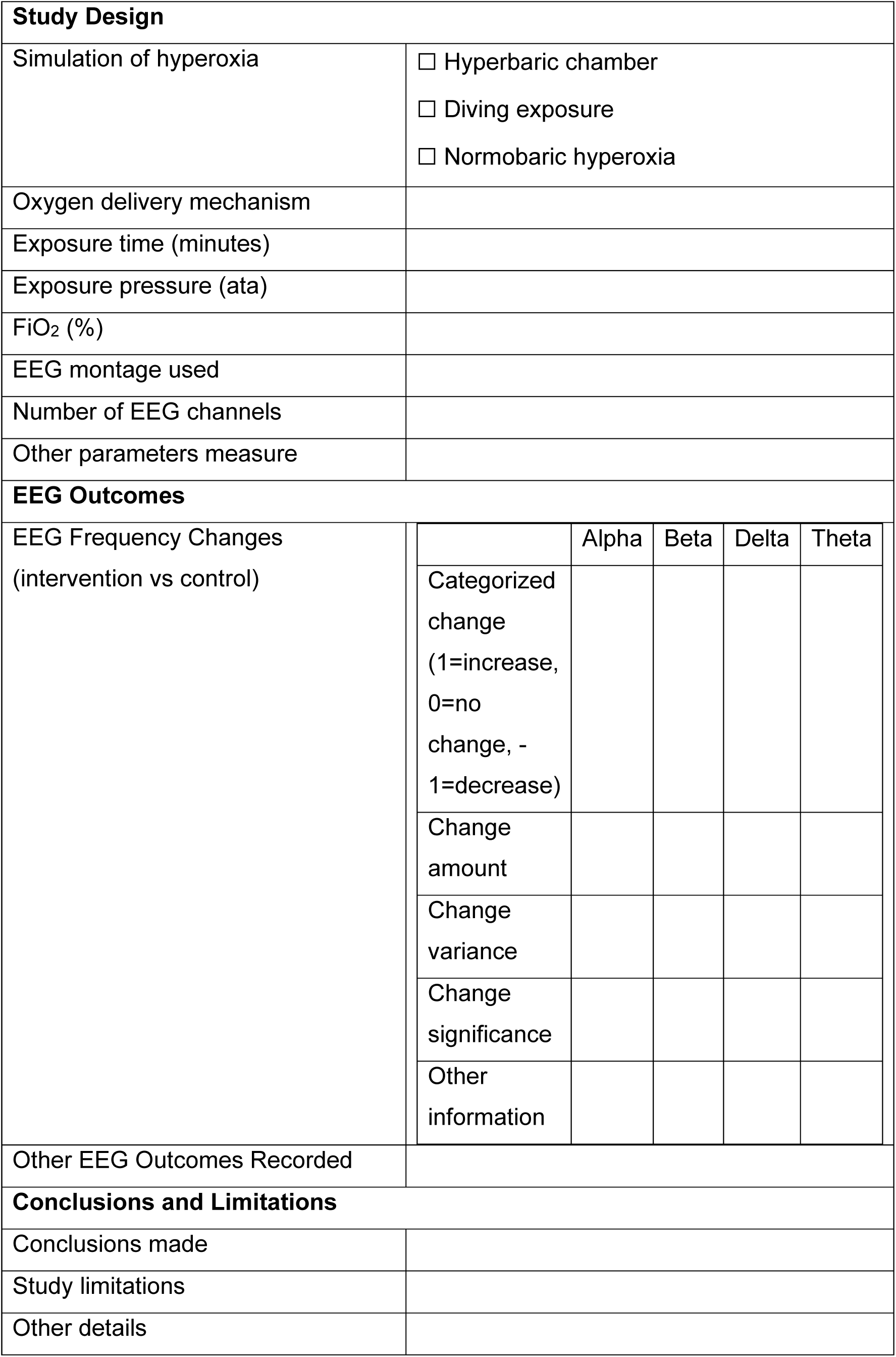

|  |  |
| --- | --- |
| **Study** |  |
| First author |  |
| Year published |  |
| Outcomes Available (pre- and post-exposure) | ☐ Frequency analysis<br>☐ Source localisation<br>☐ Connectivity analysis<br>☐ Visual expert analysis<br>☐ Evoked potentials |
| Study level of evidence | ☐ Systematic review<br>☐ Randomised control trial<br>☐ Non-randomised or non-controller trial<br>☐ Case series<br>☐ Mechanism based reasoning |
| **Participant Demographics** |  |
| Number of participants |  |
| Percentage of participants male |  |
| Participant average age |  |
| Participant age variance |  |
| Type of participants | ☐ Healthy adult volunteers<br>☐ Healthy adult volunteers with some diving experience (< 1 year)<br>☐ Healthy adult volunteers with moderate diving experience (< 3 years)<br>☐ Healthy adult volunteers with significant diving experience (3+ years) |
| Percentage of participants which had completed an oxygen tolerance test |  |

|  |  |  |  |  |  |
| --- | --- | --- | --- | --- | --- |
| **Study Design** |  |  |  |  |  |
| Simulation of hyperoxia | ☐ Hyperbaric chamber<br>☐ Diving exposure<br>☐ Normobaric hyperoxia |  |  |  |  |
| Oxygen delivery mechanism |  |  |  |  |  |
| Exposure time (minutes) |  |  |  |  |  |
| Exposure pressure (ata) |  |  |  |  |  |
| FiO<sub>2</sub> (%) |  |  |  |  |  |
| EEG montage used |  |  |  |  |  |
| Number of EEG channels |  |  |  |  |  |
| Other parameters measure |  |  |  |  |  |
| **EEG Outcomes** |  |  |  |  |  |
| EEG Frequency Changes<br>(intervention vs control) |  | Alpha | Beta | Delta | Theta |
|  | Categorized change<br>(1=increase, 0=no change, -1=decrease) |  |  |  |  |
|  | Change amount |  |  |  |  |
|  | Change variance |  |  |  |  |
|  | Change significance |  |  |  |  |
|  | Other information |  |  |  |  |
| Other EEG Outcomes Recorded |  |  |  |  |  |
| **Conclusions and Limitations** |  |  |  |  |  |
| Conclusions made |  |  |  |  |  |
| Study limitations |  |  |  |  |  |
| Other details |  |  |  |  |  |

### QUALITY ASSESSMENT TEMPLATE

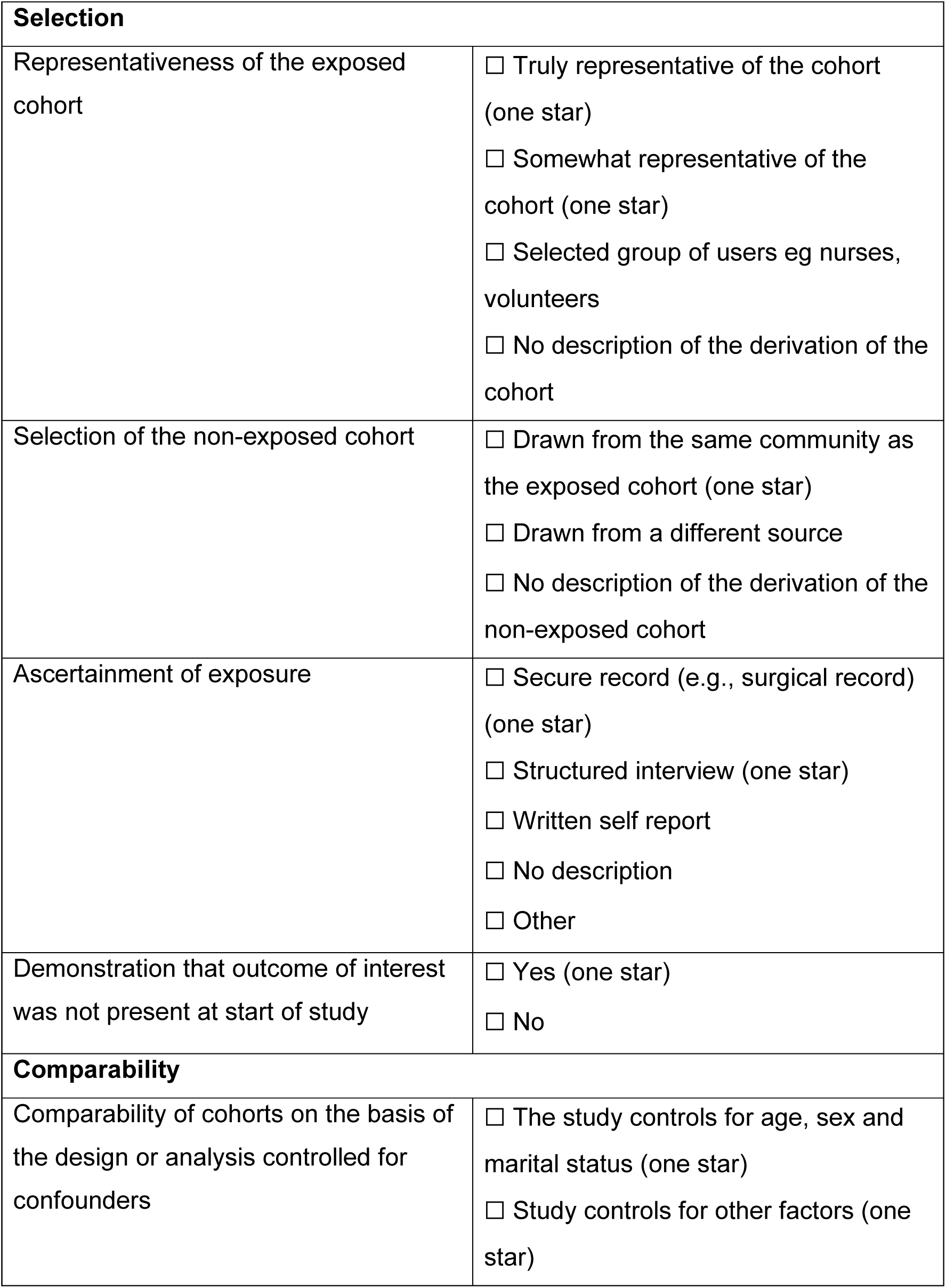
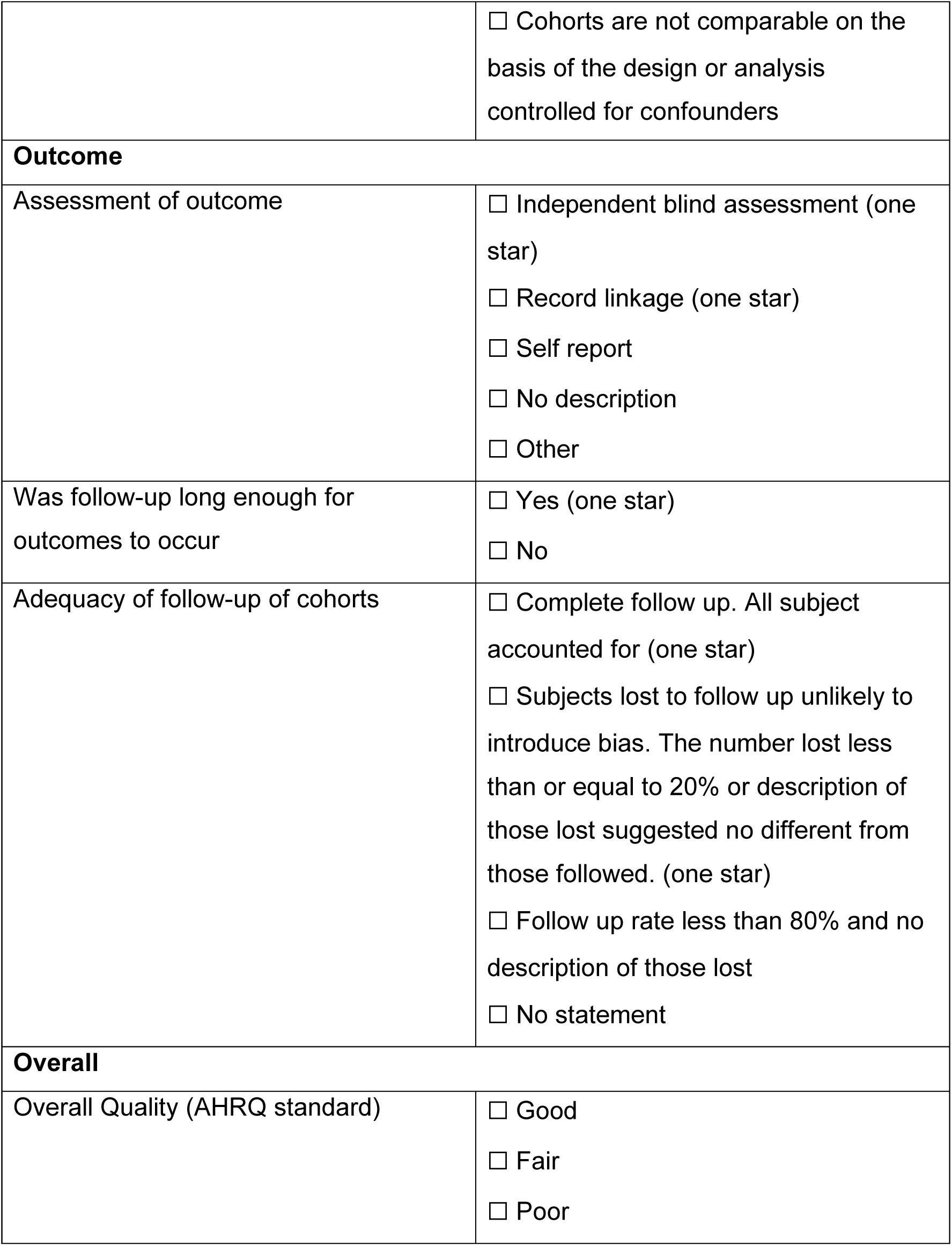

|  |  |
| --- | --- |
| **Selection** |  |
| Representativeness of the exposed cohort | ☐ Truly representative of the cohort (one star)<br>☐ Somewhat representative of the cohort (one star)<br>☐ Selected group of users eg nurses, volunteers<br>☐ No description of the derivation of the cohort |
| Selection of the non-exposed cohort | ☐ Drawn from the same community as the exposed cohort (one star)<br>☐ Drawn from a different source<br>☐ No description of the derivation of the non-exposed cohort |
| Ascertainment of exposure | ☐ Secure record (e.g., surgical record) (one star)<br>☐ Structured interview (one star)<br>☐ Written self report<br>☐ No description<br>☐ Other |
| Demonstration that outcome of interest was not present at start of study | ☐ Yes (one star)<br>☐ No |
| **Comparability** |  |
| Comparability of cohorts on the basis of the design or analysis controlled for confounders | ☐ The study controls for age, sex and marital status (one star)<br>☐ Study controls for other factors (one star) |
|  | ☐ Cohorts are not comparable on the basis of the design or analysis controlled for confounders |
| **Outcome** |  |
| Assessment of outcome | ☐ Independent blind assessment (one star)<br>☐ Record linkage (one star)<br>☐ Self report<br>☐ No description<br>☐ Other |
| Was follow-up long enough for outcomes to occur | ☐ Yes (one star)<br>☐ No |
| Adequacy of follow-up of cohorts | ☐ Complete follow up. All subject accounted for (one star)<br>☐ Subjects lost to follow up unlikely to introduce bias. The number lost less than or equal to 20% or description of those lost suggested no different from those followed. (one star)<br>☐ Follow up rate less than 80% and no description of those lost<br>☐ No statement |
| **Overall** |  |
| Overall Quality (AHRQ standard) | ☐ Good<br>☐ Fair<br>☐ Poor |

## Notes

### Competing Interest Statement

The authors have declared no competing interest.

### Funding Statement

This work was supported by funding from the Office of Naval Research of The United States Navy (N00014-23-1-2467).

### Author Declarations

University of Auckland Human Participants Ethics Committee waived ethical approval for this work.

